# AugmentA: Patient-specific Augmented Atrial model Generation Tool

**DOI:** 10.1101/2022.02.13.22270835

**Authors:** Luca Azzolin, Martin Eichenlaub, Claudia Nagel, Deborah Nairn, Jorge Sánchez, Laura Unger, Olaf Dössel, Amir Jadidi, Axel Loewe

## Abstract

Digital twins of patients’ hearts are a promising tool to assess arrhythmia vulnerability and to personalize therapy. However, the process of building personalized computational models can be challenging and requires a high level of human interaction. A pipeline to standardize the generation of a patient’s atrial digital twin from clinical data is therefore desirable. We propose a patient-specific Augmented Atria generation pipeline (AugmentA) as a highly automated framework which, starting from clinical geometrical data, provides ready-to-use atrial personalized computational models. AugmentA consists firstly of a preprocessing step applied to the input geometry. Secondly, the atrial orifices are identified and labelled using only one reference point per atrium. If the user chooses to fit a statistical shape model (SSM) to the input geometry, it is first rigidly aligned with the given mean shape before a nonrigid fitting procedure is applied. AugmentA automatically generates the fiber orientation and finds local conduction velocities by minimizing the error between the simulated and clinical local activation time (LAT) map. The pipeline was tested on a cohort of 29 patients on both segmented magnetic resonance images (MRI) and electroanatomical maps of the left atrium. Moreover, the pipeline was applied to a bi-atrial volumetric mesh derived from MRI. The pipeline robustly integrated fiber orientation and anatomical region annotations in 38.4±5.7 s. The error between insilico and clinical LAT maps was on average 12.7 ms. In conclusion, AugmentA offers an automated and comprehensive pipeline delivering atrial digital twins from clinical data in procedural time.

## I. Introduction

**A**TRIAL fibrillation (AF) is a cardiac arrhythmia characterized by uncoordinated and chaotic atrial activation affecting more than 40 million people worldwide [1]. AF is the most prevalent arrhythmia and is associated with an increased long-term risk of other cardiovascular diseases. Computational modeling provides a novel framework to assess initiation, maintenance, and progression of AF in a personalized manner [2]–[7]. Cardiac digital twins (CDT) are digital replicas of patient hearts systematically integrating clinical data that match like-for-like all available clinical observations [8], [9]. Due to their intrinsic predictive ability, CDTs are a promising tool for precision medicine and personalised treatment aiding clinical decision making and providing an efficient and cost-effective platform for testing ethically and safely innovative therapies. However, current frameworks to deliver CDT integrating both the anatomical and functional twinning phases, referring to the inference of model anatomy and electrophysiology from clinical data, are not sufficiently efficient, robust, and accurate for advanced clinical and industrial applications. Several techniques to generate different sorts of atrial models have been presented covering a range of complexity and level of detail. Personalized computational models have been developed from either imaging data, e.g. derived from computed tomography (CT) or magnetic resonance images (MRI) [4], [10]–[12], or electroanatomical maps [13]. However, a standardized pipeline for generating personalised atrial computer models from geometries derived from either of the described recording modalities remains unavailable. Lately, Razeghi et al. presented the CemrgApp open source platform for image processing to provide MRI segmentation including fibrotic tissue distribution derived from late gadolinium enhancement (LGE) intensity in a semi-automatic and userfriendly way [14]. In addition, Williams et al. published an open source platform to import, preprocess and analyze electroanatomical mapping data [15]. What is missing today is a pipeline that ingests the segmentations as provided for example by CemrgApp and the functional information as provided for example by openEP and builds a simulation-ready digital twin model. Building such model directly from clinical geometrical data remains a challenging process since imaging data can be influenced by various degrees of segmentation uncertainty [16] and electroanatomical maps can miss relevant anatomical structures (right atrium, appendage, veins). Recently, Nagel et al. presented a bi-atrial statistical shape model (SSM) covering the relevant anatomical variability required for insilico electrophysiological experiments and well generalizing to unseen geometries [17]. Moreover, SSM were shown to be a valuable tool to generate large cohorts of ready-to-use atrial models to run electrophysiological simulations [18].

Even more complicated is the functional twinning phase, where the spatio-temporal myocardial depolarisation is retrieved by clinical data possibly affected by noise and uncertainty. In particular, late gadolinium enhancement distribution and/or electroanatomical maps can be used to infer cardiac tissue characteristics and estimate conduction velocity [13], [19]–[22].

We aimed to develop and provide a highly automated pipeline to generate personalized computational models of human atria augmented with population-level a-priori knowledge (fiber orientations and region annotation) suitable for in-silico experiments. We designed the platform to enhance usability and reproducibility requiring minimal user interaction.

## II. Materials and Methods

The automated modeling pipeline for generating detailed personalized computational models of human atria is illustrated in Fig. 1. The input can be an atrial surface obtained from an electroanatomical mapping system or derived from tomographic imaging segmentation (e.g. MRI or CT). In the case of a surface with closed orifices, the pipeline proceeds with the opening of atrial orifices as described in Sec. II-B and in Sec. II-C. Next, the user can decide to proceed to fit a SSM to the target anatomy in case the user would like to augment possible missing anatomical structures or to resolve segmentation uncertainties. Provided that the quality of the original geometry is ensured, the user can directly proceed to the resampling step presented in Sec. II-H. If the desired workflow includes fitting an SSM to the target geometry, the pipeline will first label the atrial orifices as presented in Sec. II-D, automatically prealign the target mesh to the average geometry of the SSM and generate the landmarks needed for the fitting procedure as explained in Sec. II-E. Consequently, the SSM is fitted as detailed in II-F. Then, the resulting model is resampled as specified in Sec. II-H. Finally, the atrial anatomical regions are annotated and fiber orientation is computed as described in Sec. II-I. We quantitatively evaluated and compared four different methodologies to estimate conduction velocity from clinical data in Sec. II-J. The proposed highly automated atrial modeling pipeline was tested on both electroanatomical maps and MRI segmentations in a cohort of 29 patients further detailed in Sec. II-A.

**Fig. 1.**
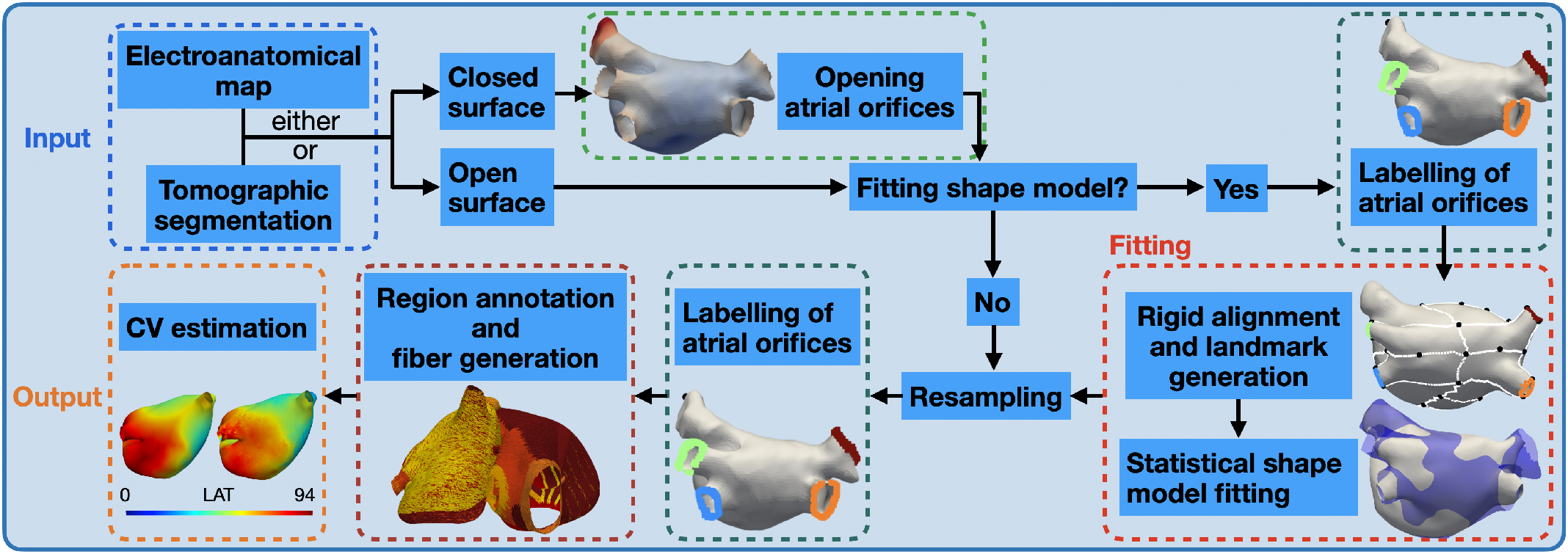
Overview of the proposed pipeline for the generation of personalised computational atrial models from different clinical anatomical data.

### A. Dataset

A cohort of 29 persistent AF patients (65±9 years, 86% male) for which both electroanatomical maps and MRI segmentations of the left atrium (LA) were available were included in this study. High density electroanatomical left atrial maps (2170± 478 sites) were acquired in sinus rhythm prior to pulmonary vein isolation using a 20-polar mapping catheter (Lasso-Nav or PentaRay-catheter, Biosense Webster Inc., CA, USA) and the CARTO 3 system (Biosense Webster Inc., CA, USA). The post-processing of the LA LGE-MRI (3 Tesla, Somatom Skyra, Siemens Healthcare, Erlangen, Germany)) was performed by an independent expert laboratory (ADAS3D Medical, Barcelona, Spain) blinded to any clinical data. The study was approved by the institutional review board, registered in the German WHO primary registry DRKS (unique identifier: DRKS00014687), and all patients provided written informed consent prior to enrollment. Moreover, a bi-atrial geometry was derived by Subject 3 presented in Krueger et al. [4]. No electroanatomical map was available in this case.

### B. Mitral valve opening annotation

For the case of a closed surface with no valve openings provided as input to the pipeline, we implemented an automated algorithm to estimate the location of the mitral valve (MV) opening. The method differs depending on whether an electroanatomical map or an MRI segmentation is supplied.

- If the input closed surface comes from an electroanatomical map, the peak-to-peak bipolar voltage was calculated from all EGM signals of the electroanatomical map in the window of the QRS complex. The QRS complex was found using the ECGdeli toolbox on the ECG lead V3 [23], [24]. Areas of high voltage (values above the 75 % voltage percentile) were considered as the MV due to the ventricular far field causing high voltages in this area. To ensure only the MV was identified and not atrial wall areas, points were only considered if they had high voltage during QRS and a low bipolar voltage (*<*0.5 mV) outside of the QRS complex.
- Since for the closed atrial surface derived from MRI, no electrogram signals are available which can be used to identify the MV, the existing mean shape of the LA where the MV was marked was used [25]. The mean shape was rigidly aligned and correspondence was established to each new patient geometry [25]. Afterwards, the area marked as MV in the mean shape could then be identified in the new geometry.

### C. Mesh pre-processing and removing valves and veins

Geometries directly derived from clinical data usually do not have a sufficient mesh quality for electrophysiological monodomain simulations [26]. Therefore, regardless of the input, AugmentA proceeds with a mesh pre-processing step, which consists in removing self-intersections and degenerate elements, while regions of the surface without defects are left unmodified. The pymeshfix Python module [27] is used in this mesh pre-processing step. Then, the veins are identified as the regions with highest surface curvature, due to their anatomical cylindrical configuration. Briefly, the surface curvature at each node is calculated as 1*/R*, where *R* is the radius of the fitted osculating sphere, as in [28]. The parameter *R* is set to 3 cm in both LA and right atrium (RA). Areas with a surface curvature higher than 1.1 times the surface curvature median value are labeled as high curvature. Then, the pipeline proceeds with opening the MV. The algorithm is different depending on whether the input geometry comes from tomographic imaging or electroanatomical mapping:

- In the case of a mesh derived from imaging data, the center of mass of the region marked as MV is computed after the co-registration presented in Sec. II-B and all the elements with a distance less than 2 cm [29] from the center of mass are removed to create the opening. Afterwards, the user has to manually select the atrial appendage apex. The pipeline continues with the clipping of the pulmonary veins by removing the elements that belong to high curvature regions and are not including the manually marked left atrial appendage apex.
- In the case of an electroanatomical map, all the elements enclosed in a sphere around the center of mass of the region marked as MV and with a diameter computed as the largest distance between all the points labelled as MV are removed. A maximum diameter of 4 cm [29] is set as upper bound to be consistent with the dimension of the MV opening from the imaging data. Occasionally, a geometrical artifact caused by the transseptal puncture is present in the electroanatomical maps. In those cases, the reference point is chosen at the tip of the transseptal puncture instead of the left atrial appendage apex. Later, the veins are identified as the areas with both high curvature and low bipolar voltage (lower than 0.5 mV), since the left atrial appendage mostly presents high voltage and veins mostly present low voltage. Each vein’s ring is generated by clipping all the elements intersecting a sphere centered at the point with maximum curvature and with variable radius chosen as the distance between the highest curvature point and the closest one with bipolar voltage higher than 0.5 mV, as shown in Fig. 3. The high curvature region manually marked as geometrical artifact due to the transseptal puncture is left out in the clipping procedure. The remaining high curvature and high voltage regions are annotated as possible left atrial appendage and the vertex with highest curvature as the apex. Following the the clipping step, the resulting processed geometry with the final openings and the automatically identified apex of the appendage is visualized using PyVista [30]. At this point, the user can decide to manually select a different vertex which is going to be used as reference point in the following labelling step.

### D. Automatic labelling of atrial orifices

The atrial orifices (pulmonary veins, MV in the LA and inferior & superior vena cava, tricuspid valve, coronary sinus in the RA) are automatically identified and labelled using a clustering algorithm presented in [31]. The only landmarks required are the apex points of the left and right atrial appendages. Briefly, the atrial orifices can be detected as the boundary edges. For the LA, the MV is determined as the largest connected boundary and the rest of the rings are clustered twice to distinguish between left and right as well as inferior and superior pulmonary veins using k-means clustering. At first, the pulmonary veins belonging to the cluster closer to the left atrial appendage (LAA) are labelled as left pulmonary veins (LPV). The remaining cluster is marked as right pulmonary veins (RPV). Secondly, the LPV are further separated into superior and inferior. The left inferior pulmonary vein is detected as the cluster closest to the LAA. The right inferior pulmonary vein is set as the one belonging to the same side as the left inferior pulmonary vein of the plane passing through the LPV, RPV and the MV centers of mass. For the orifices of the RA, the tricuspid valve (TV) is identified as the largest one. The cluster closest to the apex point of the right atrial appendage (RAA) is labelled as superior vena cava (SVC). The smallest of the remaining two rings is marked as coronary sinus (CS) and the other as inferior vena cava (IVC). Subsequently, a plane passing through the centers of the SVC, IVC and TV is used to identify a band between the IVC and the SVC and to divide the TV into a septal (*TV*_*S*_) and a lateral wall part (*TV*_*L*_). These are used as boundary conditions in the region labelling and fiber generation algorithm presented in Sec. II-I.

### E. Rigid alignment and landmark generation

After the labelling of the atrial orifices, the user can choose whether to proceed with fitting a SSM to the target geometry or directly jump to the resampling step followed by region annotation and fiber generation. In case the user wants to fit a SSM to the target geometry, a rigid alignment in space is required before the fitting procedure since translation and rotation are not represented by eigenmodes of the SSM. The transformation matrix was derived by minimizing the weighted sum of squared deviations [32] between the atrial orifices centroids of the two surfaces. 36 characteristic points were then automatically identified in the LA using geodesic paths connecting the previously marked orifices. 10 landmarks in the pulmonary veins, 4 around the MV ring, 4 in the roof, 2 in the septum, 1 in the left lateral wall, 9 in the anterior wall, 5 in the posterior wall, 1 at the LAA apex) were used for the subsequent fitting procedure. The location of the landmarks on the relative paths can be found in Sec. S.II. of the Supplementary Material.

### F. Non-rigid shape model fit using Iterative Closest Points and Gaussian Process regression

We aimed at establishing correspondence between a bi-atrial SSM [17] and the clinical geometry at hand. The geometries used in this work were not included in the process of building the SSM. Non-rigid fitting was performed by a combination of Iterative Closest Points (ICP) and Gaussian Process (GP) regression [33]. This methodology differs from the typical rigid ICP [34], which consists of identifying the best rigid transformation between two meshes. Briefly, the main steps of the classical rigid ICP are the following:

- find candidate correspondences between the moving mesh (i.e. SSM instance) and the target (i.e. clinically derived geometry) by considering the closest point on the target mesh as a candidate;
- solve for the best rigid transform between the moving mesh and the target mesh using Procrustes analysis [32];
- transform the moving mesh using this transformation;
- repeat until convergence.

The non-rigid ICP algorithm used in this study for model fitting performs the same steps. However, instead of finding a rigid transformation, it solves for a non-rigid one using GP regression [35]. The non-rigid transformation consists of the deformation that is encoded in the SSM eigenmodes. Given a set of points (in this case belonging to an instance of the SSM), we attribute the closest point on the target as a candidate correspondence to each of the points in the set. The returned sequence of points contains the candidate correspondences to the input points. The first main idea behind ICP is to use the candidate correspondences in a GP regression to find the best model instance explaining the observed deformations even though the correspondences are not perfect. We then defined a function that, given a sequence of identifiers of model points and their candidate correspondence positions, computes a GP regression based on the resulting deformation field and returns the principal component coefficients of the model instance fitting the candidate deformations best. These coefficients were then used as input to retrieve a bi-atrial geometry that fits our target LA. The non-rigid fitting of our SSM was implemented using ScalismoLab (https://scalismo.org).

### G. Co-registration of multi-modal data sets

The fitting method established correspondence between each target geometry and the SSM. Each data vector (local activation time, voltage, LGE IIR, etc.) was mapped from the original mesh to the fitted SSM using a nearest neighbour algorithm. Since the best fitted SSM instance is a deformed version of the mean shape, it always features the same number of nodes and vertex IDs. Therefore, the electrophysiological data mapped from one patient’s electroanatomical map to the resulting best fit of the SSM could be easily transferred to the respective SSM instance derived from imaging data. Since we assumed that the MRI segmentation was a better representation of the patient’s real atrial anatomy compared to the electroanatomical mesh, local activation time, uni- and bipolar voltage maps were registered to the SSM instance derived from the MRI fitting.

### H. Resampling

Even though the SSM in general already provides a good quality mesh, electrophysiological simulations performed with finite elements methods require a fine spatial resolution [26], [36]. We therefore procedeeded with a mesh resampling step providing a final high quality mesh with an average edge length of 0.4 mm. A combined smoothing and upsampling algorithm was performed using one iteration of Laplacian smoothing and the isotropic explicit remeshing filter of PyMeshLab [37], a Python library interfacing to MeshLab [38], an open source software to edit and process 3D triangular meshes.

### I. Automated region annotation and fiber generation algorithm

Atrial fiber architecture is characterized by the presence of multiple overlapping bundles running along different directions, differently from the ventricular fibers architecture where myofibers are aligned along regular patterns. Rule-based methods are widely used strategies to generate myocardial fiber orientation in computational cardiac models [39], [40]. A particular class of algorithms is known as Laplace–DirichletRule-Based methods (LDRBM) since they rely on the solution of Laplace problems with Dirichlet boundary conditions. In this work, a fully automated method to annotate the different atrial regions and for generating fiber orientation, based on the LDRBM proposed by Piersanti et al. [41] and updated in Zheng et al. [31] was implemented.

In the pipeline, six Laplace problems for the LA and six for the RA were formulated:

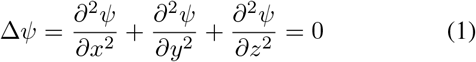

with proper Dirichlet boundary conditions *Ψ*_*a*_ and *Ψ*_*b*_ on the respective boundaries Γ_*a*_ and Γ_*b*_. These partial differential equations were solved using the open electrophysiology simulator openCARP [42]. The domain of the Laplace problems is the atrial mesh. The boundary condition domains were obtained using the method described in Sec. II-D. We further decreased the number of required manually selected seed points from four to two (apex of LAA and RAA), thus decreasing user interaction and effort and enhancing reproducibility. The two additional landmarks needed previously in [31] on the LAA basis were automatically identified by solving two additional Laplace equations *Ψ*_*ab*2_ with *Ψ*_*a*_ = 0 in the RPV and *Ψ*_*b*_ = 1 at the LAA apex and *Ψ*_*r*2_ with *Ψ*_*a*_ = 0 at both LPV and RPV and *Ψ*_*b*_ = 1 at the MV ring as Dirichlet boundary conditions. In addition, a supplementary Laplace system *Ψ*_*v*2_ with *Ψ*_*a*_ = 1 in the IVC and *Ψ*_*b*_ = 0 at the RAA apex is solved in the RA to improve the identification of the RAA compared to Zheng et al. [31]. The complete list of boundary conditions used can be found in Tab.1 of the Supplementary Material. The bundle selection was automatically performed and the bundles’ dimension was adjusted for each patient using a region growing method. Following the bundle selection, an orthonormal local coordinate system was built at each element of the atrial domain by performing a Gram–Schmidt orthogonalization. In the case that only an endocardial surface mesh is provided as input, we computed the transversal direction as the normal vector to the atrial surface at each point and the option of generating a bilayer model [43] with different endo- and epicardial fiber arrangement was included in the pipeline.

### J. Conduction velocity estimation methods

Cardiac tissue conduction velocity (CV) was estimated using four different methodologies. Regional anisotropy ratio was fixed to 3.75:1 in the LA [44]. In the first two, only non-invasive data coming from the LGE-MRI were used (i.e., IIR). The third and fourth method, estimate the CV from the clinically recorded local activation time (LAT) map.

- The first method consisted in discretely applying different CV depending on the IIR value (CV_di_). We tuned the monodomain conductivity to reach a longitudinal CV of 1.0 m/s in the healthy tissue (IIR*<*1.2), of 0.7 m/s in the interstitial fibrosis areas (1.2*≤*IIR*<*1.32) and of 0.6 m/s in the dense fibrosis regions (IIR*≥*1.32) [19], [45].
- In the second method we applied a CV following a regression model (CV_rm_) relating CV to IIR value [45]:

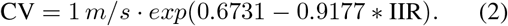

- In the third method, we calculated the CV by fitting radial basis functions (CV_rb_) [21], [46]. The algorithm selects stable catheter positions, finds the local activation times (LAT), considers the wall contact and calculates all CV estimates within the area covered by the catheter.
- The fourth method consisted in iteratively tuning (CV_tu_) each element conductivity to minimize the root mean squared error (RMSE) between the simulated LAT and the recorded clinical LAT. The fiber field used is the one presented in Sec. II-I. The clinical LAT map is divided into activation bands from the earliest to the latest activation in steps of 30 ms. However, selecting the location of earliest activation as the very first activated map point or electrogram point can be error-prone. Therefore, the earliest activated point is identified as the center of mass of the region with clinical LAT within the 2.5th percentile. Firstly, the earliest 2.5th percentile LAT points are identified and then their center of mass is calculated and used as earliest activated point in the in-silico experiment. Before proceeding with the iterative fitting of the clinical LAT, we detect the nodes with an earlier activation than the neighboring vertices and mark them as wrong annotations, as illustrated in Fig. 9B. The LAT annotations in these areas are not used in the fitting process and the conductivity is chosen as the mean of the region boundary.

### K. Atrial models and computational tools

The myocyte membrane dynamics were represented with a variant of the original Courtemanche et al. model [47] reflecting AF-induced remodeling [48]. Atrial tissue regions with IIR higher than 1.2 were labelled as fibrosis. We set 30% of the elements in the fibrotic regions as almost not conductive (conductivity of 10^*−*7^ S/m) to account for structural remodeling and the presence of scar tissue. This approach modelled the macroscopic passive barrier behaviour caused by the electrical decoupling of the myocytes in the tissue infiltrated by fibrosis, also referred to as ‘percolation’ [49]. In the other 70%, several ionic conductances were rescaled to consider effects of cytokine-related remodeling [50] (−50% g_*K*1_, -40% g_*Na*_ and -50% g_*CaL*_). The spread of the electrical depolarization in the atrial myocardium was simulated by solving the monodomain equation using openCARP [42] and a time step of 0.02 ms.

## III. Results

### A. Removing mitral valve and pulmonary veins

The automatic annotation of the MV opening region identified the location of the MV in all 29 patients both in electroanatomical maps and MRI segmentations. Fig. 2 shows an example of both the MV labelling step and the consequent creation of the valve ring. Regions with surface curvature higher than 1.1 times the median value identified both veins and appendage in the case of MRI segmentation and the pipeline correctly proceeded with the clipping of the veins regions, though maintaining the appendage, as shown in Fig. 3. When the input geometry was derived from an electroanatomical map, the combination of high surface curvature and low bipolar voltage resulted in an accurate location of the pulmonary vein regions in the LA. The remaining high curvature area always corresponded to the LAA.

**Fig. 2.**
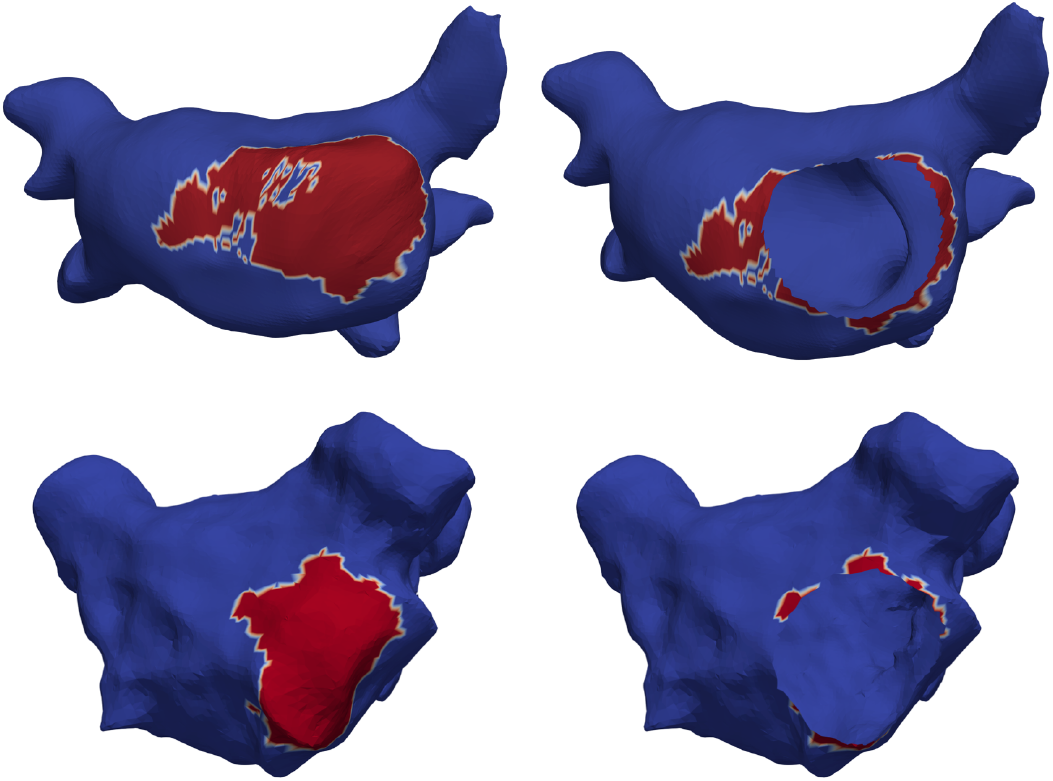
Identification and generation of mitral valve opening. Elements marked as mitral valve (in red) after the co-registration procedure and the resulting mitral valve opening. The remaining high curvature region is the left atrial appendage. Top row: example based on an MRI segmentation (patient 28). Bottom row: example based on an electroanatomical map (patient 28).

**Fig. 3.**
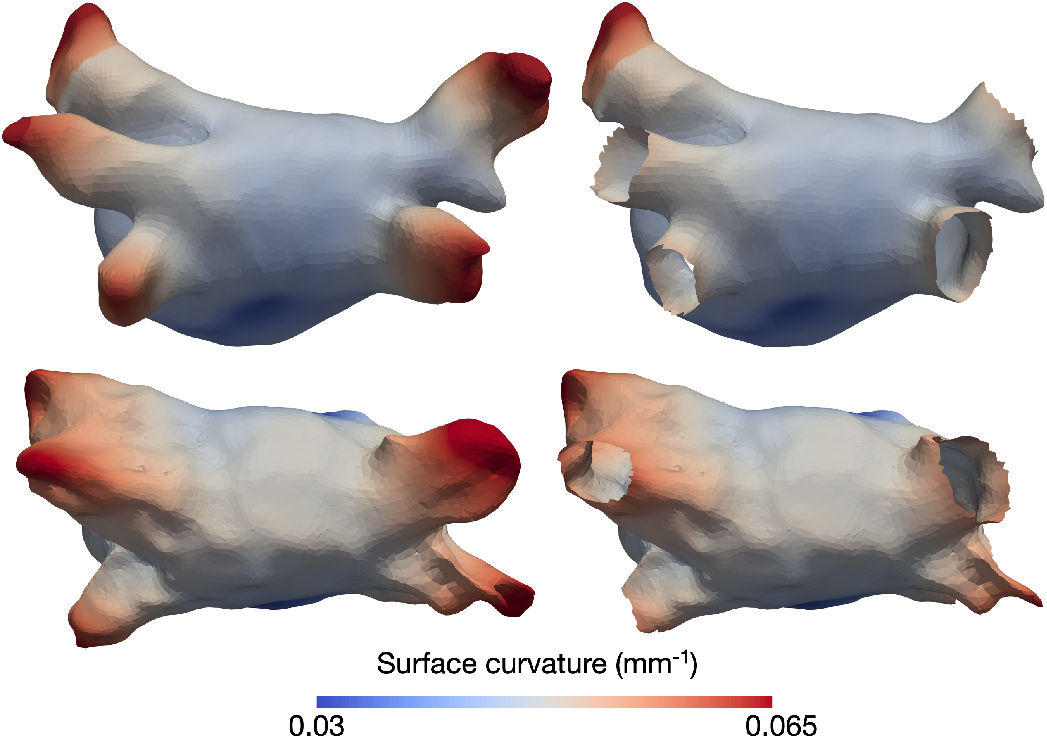
Pulmonary veins clipping using the surface curvature. Regions with surface curvature higher than 1.1 the median value are shown in red and the mesh after the removal of the pulmonary veins in the right column. Top row: example based on an MRI segmentation (patient 28). Bottom row: example based on an electroanatomical map (patient 28).

### B. Atrial orifices labelling

Atrial orifices were automatically identified and correctly labelled from the pipeline in all 29 patients. The accuracy of the labelling was checked by visual inspection. An example of the resulting annotated geometry in both an MRI segmentation and an electroanatomical map is shown in Fig. 4. The decision of using only the atrial appendage apexes as reference point for the labelling method turned out to be appropriate to robustly discriminate between valves and veins in both left and right atrium.

**Fig. 4.**
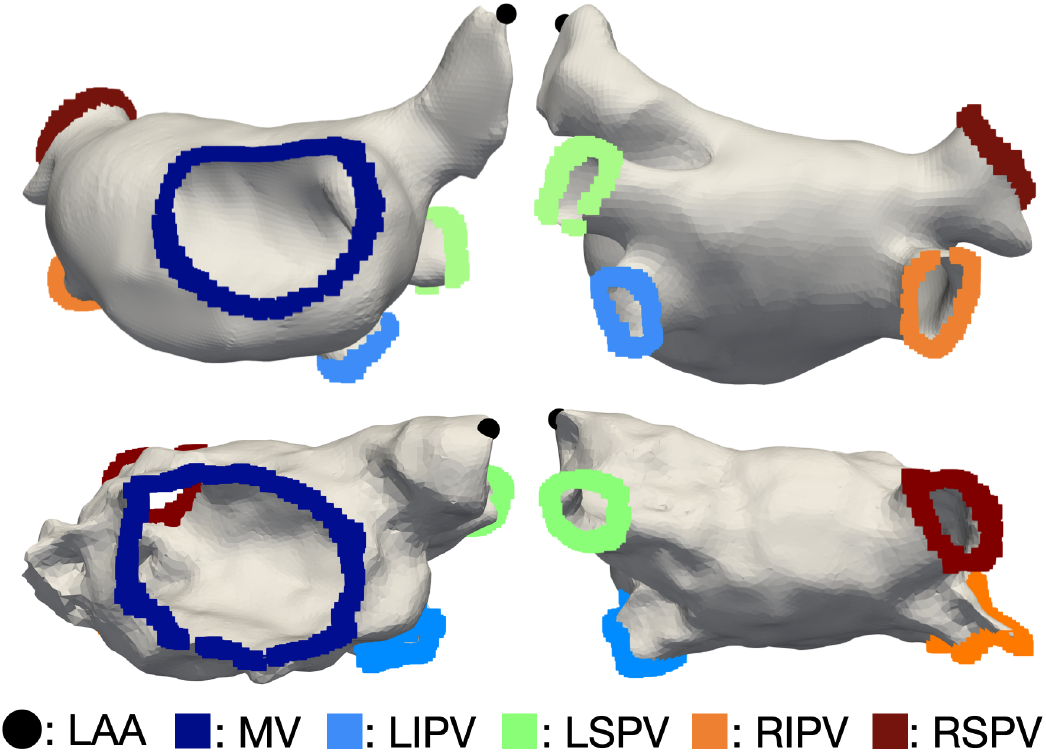
Output of the atrial openings annotation step (mitral valve and roof view of the labelled atrial orifices). Top row: example based on an MRI segmentation (patient 28). Bottom row: example based on an electroanatomical map (patient 28). LAA: left atrial appendage apex, MV: mitral valve, LIPV: left inferior pulmonary vein, LSPV: left superior pulmonary vein, RIPV: right inferior pulmonary vein, RSPV: right superior pulmonary vein.

### C. Statistical shape model fitting process

All 36 landmarks were correctly and automatically identified in the whole patient cohort using the geodesic paths connecting the atrial orifices labelled in the previous step. The reference points covered most of the atrial surface and localized the most important atrial structures, as shown in Fig. 5. The non-rigid fitting procedure provided the best fitted SSM instance for each electroanatomical map and MRI segmentation, as presented in Fig. 6. Fig. 7 shows the surface-to-surface distance between the best fitted SSM and the target electroanatomical map (2.72± 2.17 mm). The surface-to-surface distance in the case of target atrial geometries derived from MRI segmentations was 2.13± 1.79 mm across all patients (Fig. 7).

**Fig. 5.**
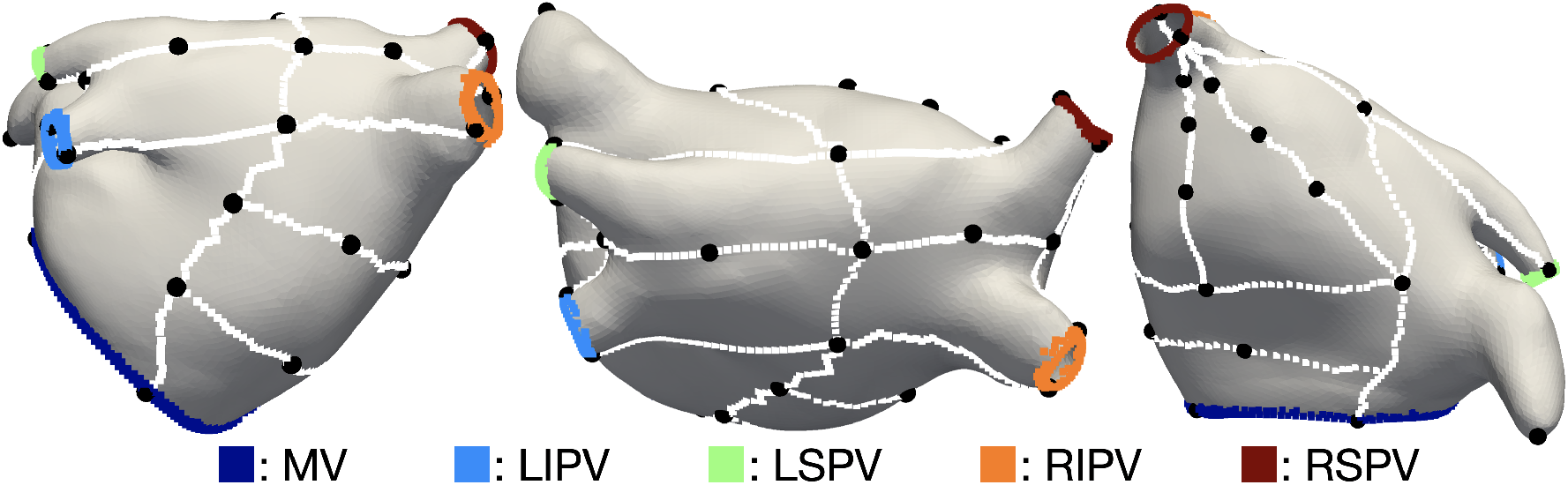
Posterior, roof and anterior view of the landmarks generation procedure on the left SSM mean instance [17]. Geodesic paths used to identify the landmarks locations in white and respective 36 landmarks in black.

**Fig. 6.**
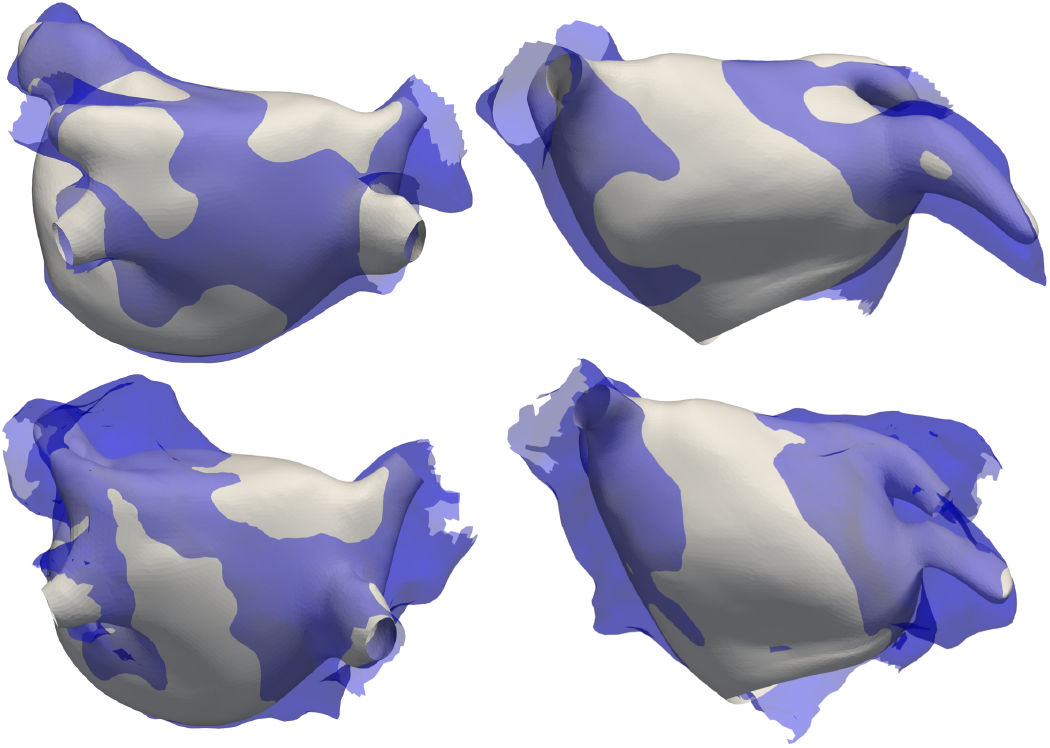
Top row: Roof and anterior view of the SSM instance (grey) best fitting to the target MRI segmentation (semi-transparent blue). Bottom row: Roof and anterior view of the SSM instance (grey) best fitting to the target electroanatomical map (semi-transparent blue).

**Fig. 7.**
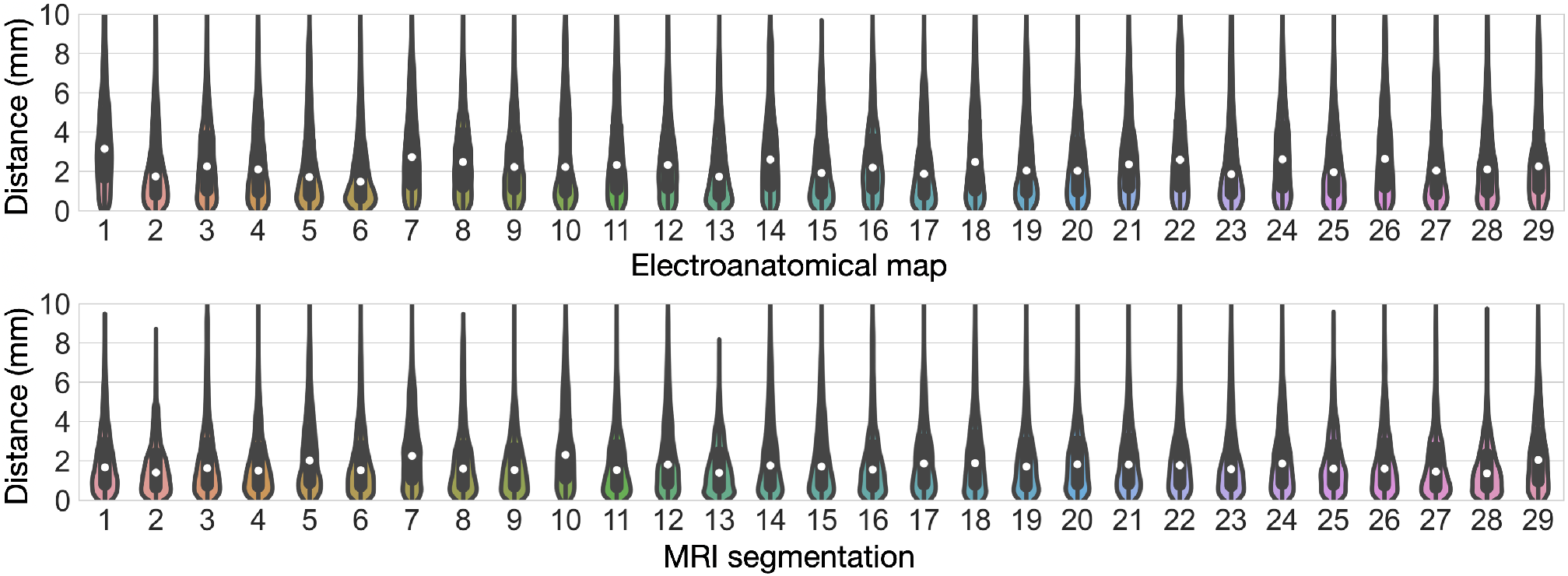
Top row: surface-to-surface distance between the best fitting SSM instance and the respective electroanatomical map. Bottom row: surface-to-surface distance between the best fitting SSM instance and the respective MRI segmentation.

### D. Atrial region annotation and fiber orientation generation

Atrial structures were automatically annotated in all 29 LA SSM instances and the pipeline carefully identified each region using the previously labelled atrial orifices and the LAA apex. Finally, bilayer models including realistic fiber orientation were generated. The pipeline was therefore tested on a bi-atrial surface model coming from the mean shape of a SSM [17]. In both LA and RA, regions were correctly annotated (Fig. 8). Moreover, fiber orientation was calculated and a bi-atrial bilayer model was generated along with interatrial bundles, as presented in Fig. 8. We recall that in the case of a bi-atrial surface as input, the only two manual reference points needed by the full pipeline are left and right atrial appendage apexes. In Fig. 8, we show the results of region annotation and fiber orientation on a volumetric bi-atrial mesh derived from MRI segmentation [4]. The same pipeline was further applied to 100 different volumetric atrial models with homogeneous thickness coming from various instances of a SSM [17] and made publicly available [51]. In Zheng et al. [31], a volumetric bi-atrial mesh with heterogeneous myocardial thickness [52] was used as input and the pipeline proceeded with region annotation and fiber generation.

**Fig. 8.**
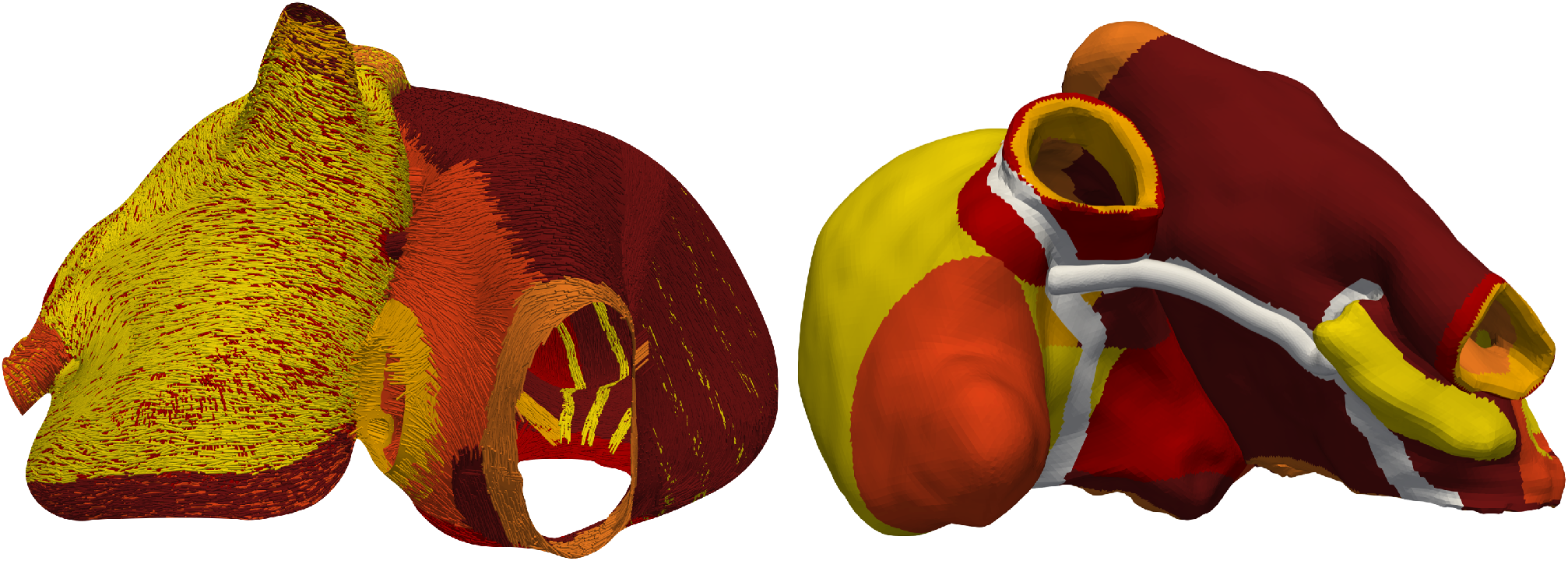
Posterior view of the fiber orientation computed with AugmentA in a bi-atrial bilayer model derived from a SSM [17] and anterior view of the region annotation in a bi-atrial volumetric model [4] in which the wide Bachmann’s Bundle is highlighted in white.

### E. Conduction velocity estimation

The simulated LAT map based on an underlying CV as estimated with the fourth method from the clinical LAT for patient 1 is shown in Fig. 9. The in-silico LAT maps computed with all four CV estimation methodologies presented in Sec. II-J can be found in Fig. S1 of the Supplementary Material. The root mean square error (RMSE) between the simulated and the clinical LAT maps was calculated for each of the presented CV estimation method and presented in Tab. I.

**Fig. 9.**
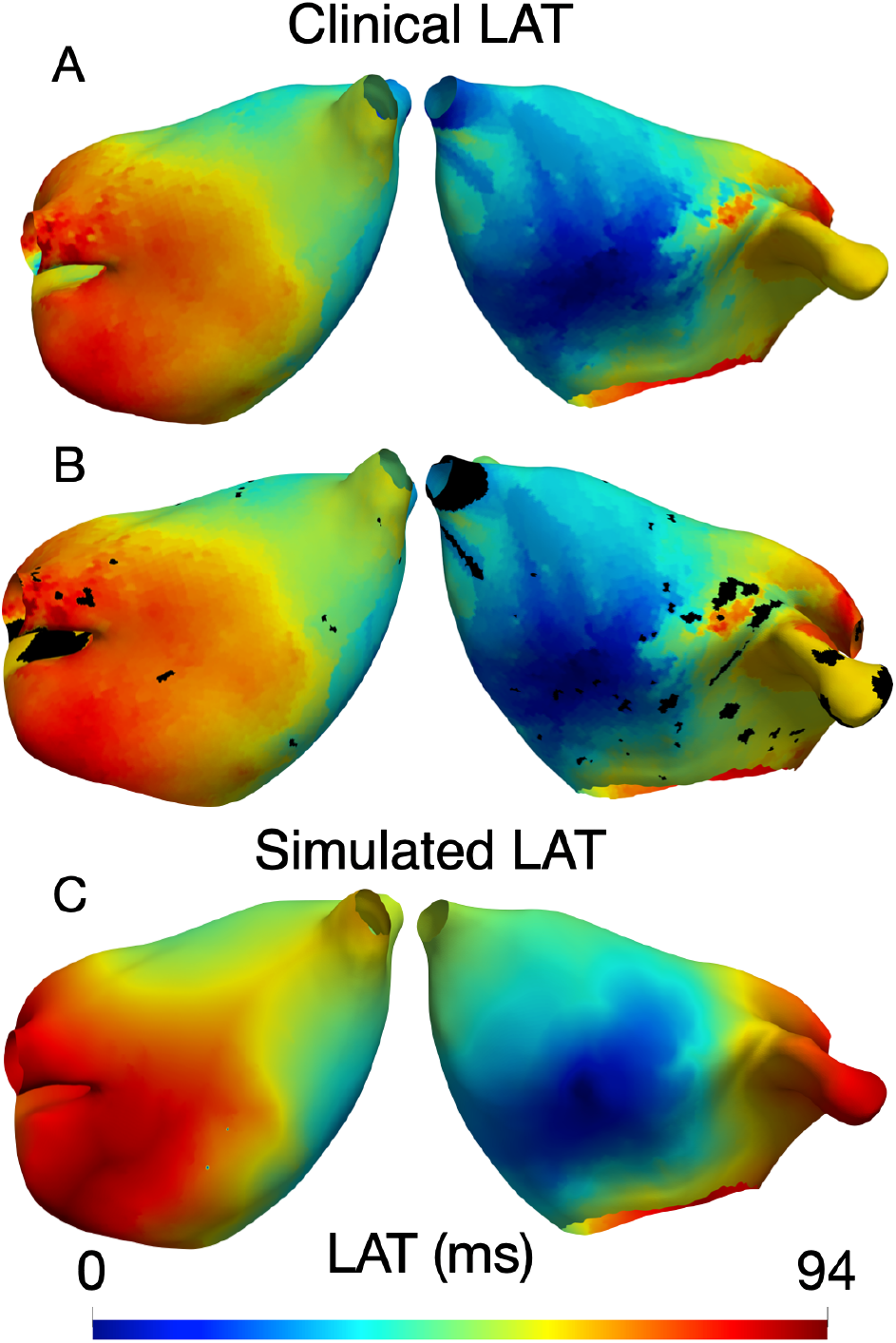
A: posterior and anterior view of the clinical LAT map of patient 1. B: posterior and anterior view of the clinical LAT map of patient 1 in which we marked in black the nodes with an earlier activation with respect to the neighbours. C: posterior and anterior view of the simulated LAT map of patient 1 using the CV_tu_ estimation method.

**TABLE I.** RMSE (ms) between the simulated and the clinical LAT maps using the different cv estimation methods.

| CV estimation method | RMSE (ms) |
| --- | --- |
| CV<sub>di</sub> | 27.1±13.6 |
| CV<sub>rm</sub> | 36.7±14.1 |
| CV<sub>rb</sub> | 107.5±44.5 |
| CV<sub>tu</sub> | 12.7±5.8 |

Moreover, we computed the relative error per patient between the simulated and the clinical LAT map as:

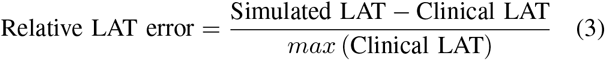

and the distributions of the relative LAT error using the various methodologies to estimate CV are shown in Fig. 10. The relative LAT errors for each patient can be found in Fig.S2-S5 the Supplementary Material.

**Fig. 10.**
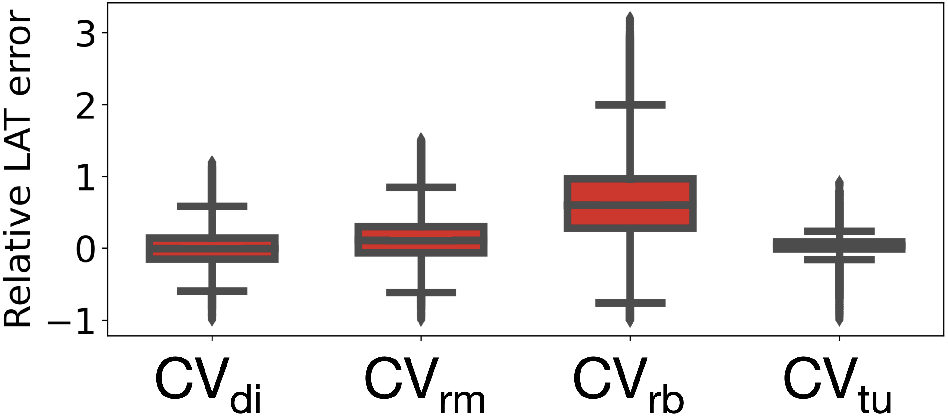
Relative LAT error between simulated and clinical LAT map using the four conduction velocity estimation methods.

### F. Processing time

The full pipeline took 38.4±5.7 s to complete the opening and labelling of atrial orifices, the fitting process, mesh resampling, the region annotation and fiber generation steps, demonstrating the feasibility of an in-situ application.

## IV. Discussion

### A. Opening and labelling atrial orifices

The pipeline automatically identified and proceeded with the opening of the atrial orifices. In comparison to CemrgApp [14], in which the user has to manually select the mitral valve and all pulmonary vein regions, the only user-interaction needed by our pipeline is the selection of the atrial appendage apex.

### B. Statistical shape model fitting

The SSM fitting step allowed to establish correspondences between each geometry derived from clinical data and the SSM. Regions with higher distance were located mostly close to the appendage. However, we have to recall that the LAA volume is often only partly covered during electroanatomical mapping. The average distance of 2.13 mm between the original MRI segmentation is in the range of one to two voxels [17], [53] omitting segmentation uncertainty. Therefore, the best fitted SSM instance very well represented the original anatomy and augmented the missing anatomical structures (e.g. appendages), as shown in our previous work [33]. The SSM generalized well to unseen geometries and accurately represented relevant anatomical features. Moreover, the patient-specific generalization in regions with high inter-individual variability such as the appendages and the PVs could be improved by updating the SSM with more clinical data.

### C. Region annotation and fiber generation

We have developed a highly automated pipeline that accurately generated the fiber direction of the human atria according to histological data. Additionally, we provide a pipeline that annotates the anatomical regions independently from the atrial geometrical variability due to the region growing implementation. We demonstrated how the pipeline can generate both volumetric and bilayer 3D models, perform with anatomical variations, and create accurate models of the human atrial anatomy. The pipeline well represented the complex atrial fiber architecture and captured the complex fiber bundles arrangements in both LA and RA. Different endocardial and epicardial fiber orientation were included to faithfully represent the transmural variability even in a bilayer computational model. Comparing to the existing method by Wachter et al. [40] that requires 21 manually defined seed points and a longer calculation time, our pipeline significantly reduces the calculation time and human manual interaction. The original LDRBM by Piersanti et al. [41] and updated by Zheng et al. [31] was further improved enhancing automation and reproducibility by limiting the manual user interaction to the selection of one reference point per atrium and using an extra set of Laplace solutions. Roney et al. [52] presented a methodology to assign the fiber orientation by mapping the fiber field from a human atrial fiber atlas to different patient specific atrial models. Even if the impact of the fiber field on average activation times computed in paced rhythm was relatively small, it had a larger effect on maximum LAT differences. Moreover, arrhythmia dynamics were highly dependent on the fiber field, suggesting that atrial fiber fields should be carefully assigned to patient-specific arrhythmia models. Our pipeline ensures personalization of both region annotation and specification of fiber orientation to each anatomical structure in the human atria.

### D. Conduction velocity estimation

We systematically evaluated three state-of-the-art methods to estimate cardiac tissue CV from clinical data and compared them to our newly proposed iterative fitting to the clinical LAT map algorithm. Monodomain conductivity in each element was calculated directly from the CV map using a regression curve representing the relationship between CV and conductivity computed tuning the tissue conductivities to match a specific CV in a tissue slab. This approach can potentially underestimate longitudinal CV if the excitation direction is not completely parallel to the myocyte orientation. This might be part of the reason for the bias towards positive errors. The CV_rb_ method had the highest RMSE due to its sensitivity to sparse data recordings.

## V. Limitations

The used SSM did not include different numbers of PVs. Therefore, the fitted SSM instance will always provide an atrial surface with four pulmonary veins. In the multi-modal data co-registration step, the data vector interpolation rather than nearest neighbor mapping might improve data fidelity.

## Data Availability

All personalized computational models used in this study including fiber orientation and anatomical labels are available online (https://doi.org/10.5281/zenodo.5589289).

## VII. Conclusion

The pipeline presented in this work offers a highly automated, comprehensive and reproducible framework to process geometries derived from clinical data and generate atrial anatomical and functional digital twins. AugmentA was able to provide ready-to-use atrial models starting from either electroanatomical maps or imaging data. The pipeline incorporates various region labels and detailed fiber orientation reducing the user-interaction to the selection of one reference point per atrium. The pipeline was moreover tested with surface and volumetric input and it correctly proceeded with atrial structure annotation and fiber generation. AugmentA could thus become a default framework for the generation of atrial digital twin models from clinical data in future studies. This work is a step forward to improve comparability and reproducibility of atrial models derived from clinical data and to facilitate the evaluation of arrhythmia vulnerability and ablation planning automating the computational model generation steps.

