## Supplementary Material for "AugmentA: Patient-specific Augmented Atrial model Generation Tool"

TABLE I
DIRICHLET BOUNDARY CONDITIONS OF THE TWELVE LAPLACE PROBLEMS.

| Atrium | $\psi$ | $\psi_a$ | $\Gamma_a$ | $\psi_b$ | $\Gamma_b$ |
|---|---|---|---|---|---|
| LA | $\psi_{tran}$ | 1 | $\Gamma_{epi}$ | 0 | $\Gamma_{endo}$ |
| | $\psi_{ab}$ | 2 | $\Gamma_{rpv}$ | 1 | $\Gamma_{mv}$ |
| | | 0 | $\Gamma_{lpv}$ | -1 | $\Gamma_{ap}$ |
| | $\psi_{ab2}$ | 1 | $\Gamma_{ap}$ | 0 | $\Gamma_{rpv}$ |
| | $\psi_v$ | 1 | $\Gamma_{rpv}$ | 0 | $\Gamma_{lpv}$ |
| | $\psi_r$ | 1 | $\Gamma_{mv}$ | 0 | $\Gamma_{lpv} \cup \Gamma_{rpv} \cup \Gamma_{mv}$ |
| | $\psi_{r2}$ | 1 | $\Gamma_{mv}$ | 0 | $\Gamma_{rpv} \cup \Gamma_{lpv}$ |
| RA | $\psi_{tran}$ | 1 | $\Gamma_{epi}$ | 0 | $\Gamma_{endo}$ |
| | $\psi_{ab}$ | 2 | $\Gamma_{ivc}$ | 1 | $\Gamma_{tv}$ |
| | | 0 | $\Gamma_{svc}$ | -1 | $\Gamma_{ap}$ |
| | $\psi_v$ | 1 | $\Gamma_{ivc}$ | 0 | $\Gamma_{svc} \cup \Gamma_{ap}$ |
| | $\psi_{v2}$ | 1 | $\Gamma_{ivc}$ | 0 | $\Gamma_{ap}$ |
| | $\psi_r$ | 1 | $\Gamma_{tv}$ | 0 | $\Gamma_{top}$ |
| | $\psi_w$ | 1 | $\Gamma_{tv\_s}$ | 0 | $\Gamma_{tv\_l}$ |

### S.I. Fiber generation

The Dirichlet boundary for the twelve Laplace problem are presented in Tab. I, in which the values $\psi_a$ and $\psi_b$ are applied on the boundary regions $\Gamma_a$ and $\Gamma_b$.

### S.II. Landmarks

1) ls_o: point on left superior PV farthest away from left inferior PV;
2) ls_i: point on left superior PV closest to left inferior PV;
3) li_i: point on left superior PV closest to left inferior PV;
4) li_o point on left inferior PV farthest away from left superior PV;
5) rs_o: point on right superior PV farthest away from right inferior PV;
6) rs_i: point on right superior PV closest to right inferior PV;
7) ri_i: point on right superior PV closest to right inferior PV;
8) ri_o: point on right inferior PV farthest away from right superior PV;
9) mv_l: point on the left lateral side of the mitral valve ring;
10) mv_r: point on the septal side of the mitral valve ring;
11) mv_an_middle: point on the anterior wall of the mitral valve ring;
12) mv_po_middle: point on the septal side of the mitral valve ring;
13) lpv_base: point at the left pulmonary veins base;
14) roof_25: point at 25% of the path on the roof connecting lpv_base to rpv_base;
15) roof_50: point at 50% of the path on the roof connecting lpv_base to rpv_base;
16) roof_75: point at 75% of the path on the roof connecting lpv_base to rpv_base;
17) rpv_base: point at the left pulmonary veins base;
18) lat_50: point at 50% of the path on the left lateral wall connecting lpv_base to mv_l;
19) laa: left atrial appendage apex;
20) sep_50: point at 50% of the path on the septal wall connecting rpv_base to mv_r;
21) sep_25: point at 25% of the path on the septal wall connecting rpv_base to mv_r;
22) p1: point at 50% of the path on the roof connecting ls_o to rs_o;
23) p1_mv_50: point at 50% of the path connecting p1 to mv_an_middle;
24) p1_mv_sep_50: point at 50% of the path connecting p1_mv_50 to sep_50;
25) p1_mv_sep_75: point at 50% of the path connecting p1_mv_75(point at 75% of the path connecting p1 to mv_an_middle) to sep_25;
26) p2: point at 50% of the path on the roof connecting li_o to ri_o;
27) p2_mv_33: point at 33% of the path connecting p2 to mv_po_middle;
28) p2_mv_66: point at 66% of the path connecting p2 to mv_po_middle;
29) p2_mv_sep_50: point at 50% of the path connecting p2_mv_33 to sep_50;
30) p2_mv_sep_75: point at 75% of the path connecting p2_mv_66 to sep_25;
31) rspvo_p1_mv_30: point at 30% of the path connecting rs_o to p1_mv_50;
32) rspvo_p1_mv_50: point at 50% of the path connecting rs_o to p1_mv_50;
33) rspvo_p1_mv_70: point at 70% of the path connecting rs_o to p1_mv_50;
34) rspvo_p1_mv_sep_30: point at 30% of the path connect-

ing rs_o to p1_mv_sep_50;

35) rspvo_p1_mv_sep_50: point at 50% of the path connecting rs_o to p1_mv_sep_50;

36) rspvo_p1_mv_sep_70: point at 70% of the path connecting rs_o to p1_mv_sep_50.

### S.III. Conduction velocity estimation

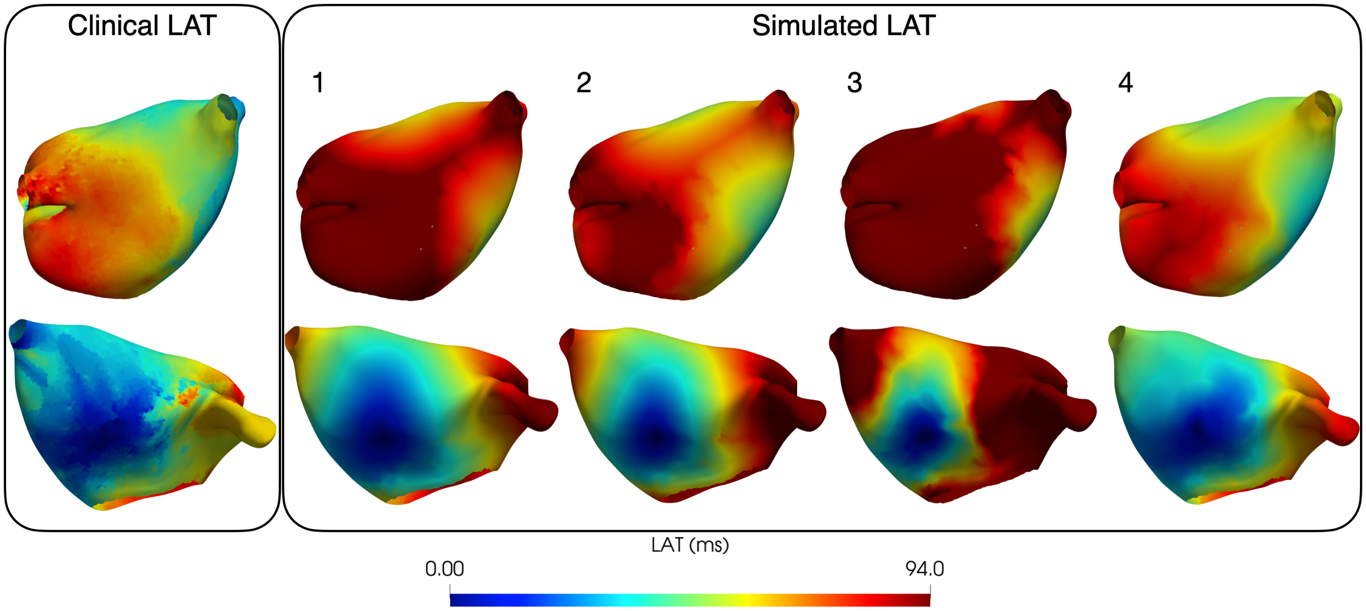


S1. First column: posterior and anterior view of the clinical LAT map of patient 1. Second column: posterior and anterior view of the patient 1 simulated LAT map using the $CV_{di}$ estimation method. Third column: posterior and anterior view of the patient 1 simulated LAT map using the $CV_{rm}$ estimation method. Fourth column: posterior and anterior view of the patient 1 simulated LAT map using the $CV_{rb}$ estimation method. Fifth column: posterior and anterior view of the patient 1 simulated LAT map using the $CV_{tu}$ estimation method.

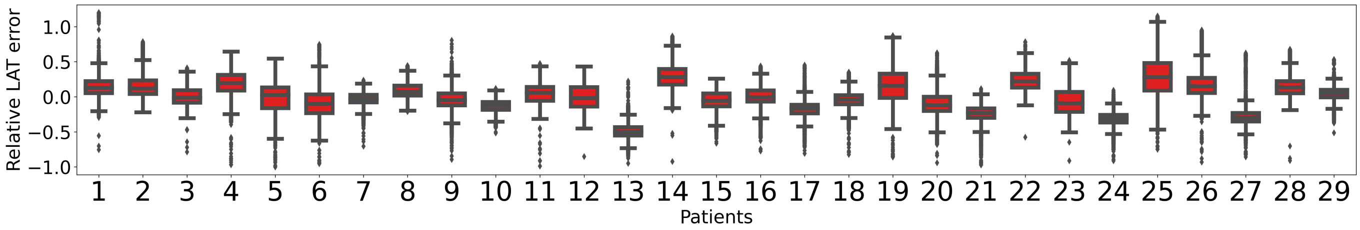


S2. Relative LAT errors per patient using the $CV_{di}$ estimation method.

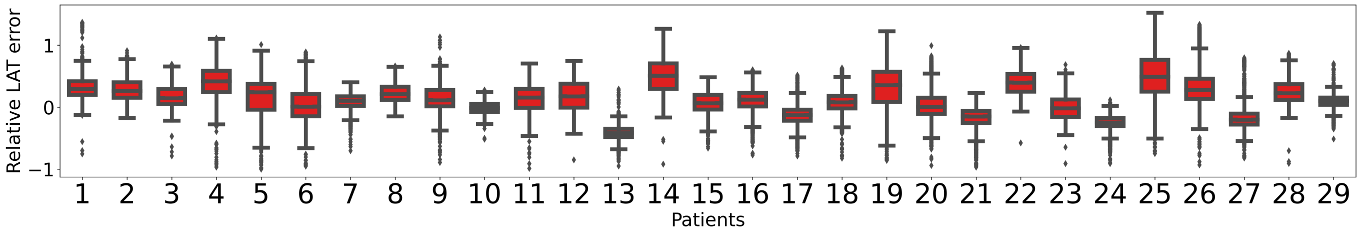


S3. Relative LAT errors per patient using the $CV_{rm}$ estimation method.

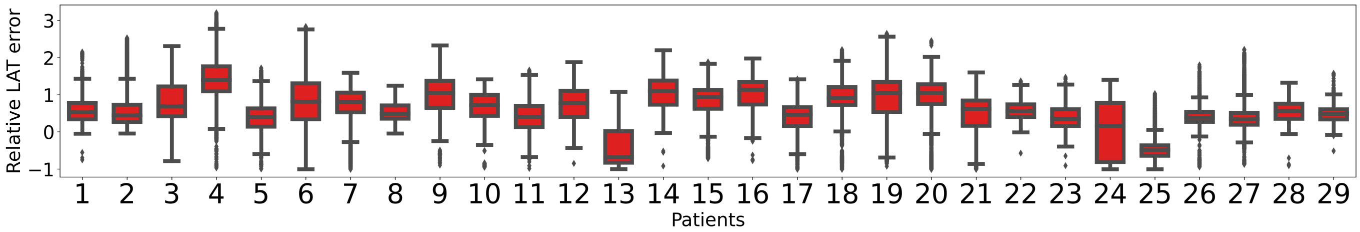


S4. Relative LAT errors per patient using the $CV_{rb}$ estimation method.

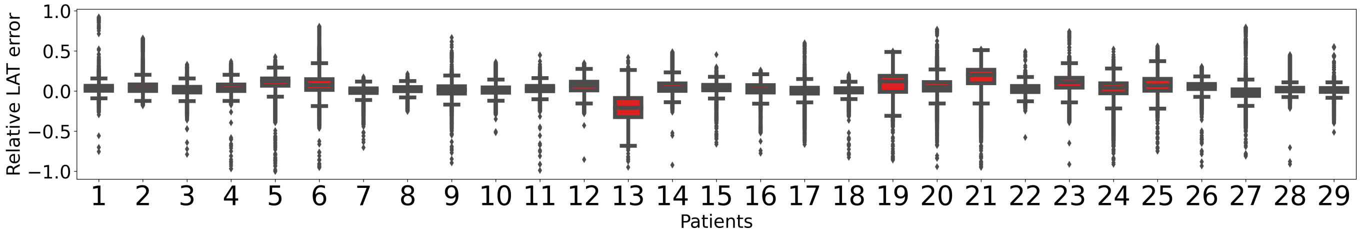


S5. Relative LAT errors per patient using the $CV_{tu}$ estimation method.
